# Evaluation of “test to return” after COVID-19 diagnosis in a Massachusetts public school district

**DOI:** 10.1101/2022.02.11.22270843

**Authors:** Sandra B. Nelson, Isaac Ravi Brenner, Elizabeth Homan, Sarah Bott Lee, Christine Bongiorno, Nira R. Pollock, Andrea Ciaranello

**Author notes:** **Address correspondence to:** Sandra B. Nelson, MD, Division of Infectious Diseases, Massachusetts General Hospital, Cox building, 5^th^ floor, Boston, MA 02114.

## Abstract

The Centers for Disease Control allows rapid antigen testing (RAT) towards the end of a 5-day isolation for COVID-19 infection to determine eligibility to leave isolation. The impact of a test-to-return (TTR) program in schools is unknown. In January 2022 a Massachusetts school district initiated a TTR program utilizing a single school-administered RAT on days 5-9 after symptom onset or positive test, whichever was first. Of 636 students with COVID-19 infection, 408 (64.2%) participated in TTR; of these, 128 (31.4%) had a positive TTR rapid antigen test. Students who were symptomatic at any time during their infection were more likely to have a positive TTR than those who were never symptomatic (43.1% vs. 17.3%); positivity rates were lower when TTR was performed later during days 6-9. TTR may identify students who carry higher viral loads after recovery from COVID-19 infection thereby extending their isolation, while facilitating earlier return of those with negative results.

## Introduction

In December 2021, the US Centers for Disease Control and Prevention (CDC) updated guidance for people with SARS-CoV-2 infection, enabling end of isolation on day 6 after symptom onset (or positive test, if asymptomatic) for those with symptom improvement and ability to mask through day 10.^1^ However, some individuals still carry culturable virus beyond day 5.^2^ Rapid antigen test (RAT) positivity correlates with higher SARS-CoV-2 viral load and the ability to culture replication-competent virus, ^3,4^ and has been considered a proxy for contagiousness.

In January 2022, CDC updated its guidance to allow RAT towards the end of the 5-day isolation. A negative test permits cessation of isolation, while a positive result extends isolation through day 10. The impact of utilizing a “test-to-return” (TTR) program in school districts is unknown.

## Methods

In January 2022, a public school district in eastern Massachusetts initiated a TTR program. Students and staff isolating for COVID-19 infection could return on day 6-10 if afebrile without antipyretics, with symptom improvement, and with a negative RAT conducted by school staff. Participants with a positive RAT and those who declined TTR maintained isolation until day 11. Eligible individuals could take a school-administered test once between days 5-9.

Data from all SARS-CoV-2 cases reported to the district with onset between December 27, 2021 and January 24, 2022 were included. Staff or student, grade level, presence of symptoms at any time during illness, date of symptom onset, date and type of initial positive test, vaccination status, and date and result of school-administered RAT for TTR participants were recorded. Day 0 of infection was considered the date of symptom onset if symptomatic or date of initial positive test, whichever came first. Eligibility for TTR and return to school was calculated forward from day 0.

Positivity rates were calculated for the entire cohort and compared by grade level, type of initial test, vaccination status, presence of symptoms during illness, and day of infection on which TTR was conducted (chi square tests, R software).^5^ Data among staff cases were insufficient for analysis.

## Results

Cases were reported among 636 students (Table 1); 67.8% with known symptom status were symptomatic. Of 408 TTR participants (64.2% of total), 128 (31.4%) had a positive RAT. There were no differences in TTR positivity by grade level, type of initial test, or vaccination status. Compared to always-asymptomatic students, ever-symptomatic students were more likely to have a positive TTR RAT (75/174[43.1%] vs. 18/104[17.3%]). TTR positivity decreased by day of infection (p=0.02; Table 1 and Figure 1).

**Table 1.**
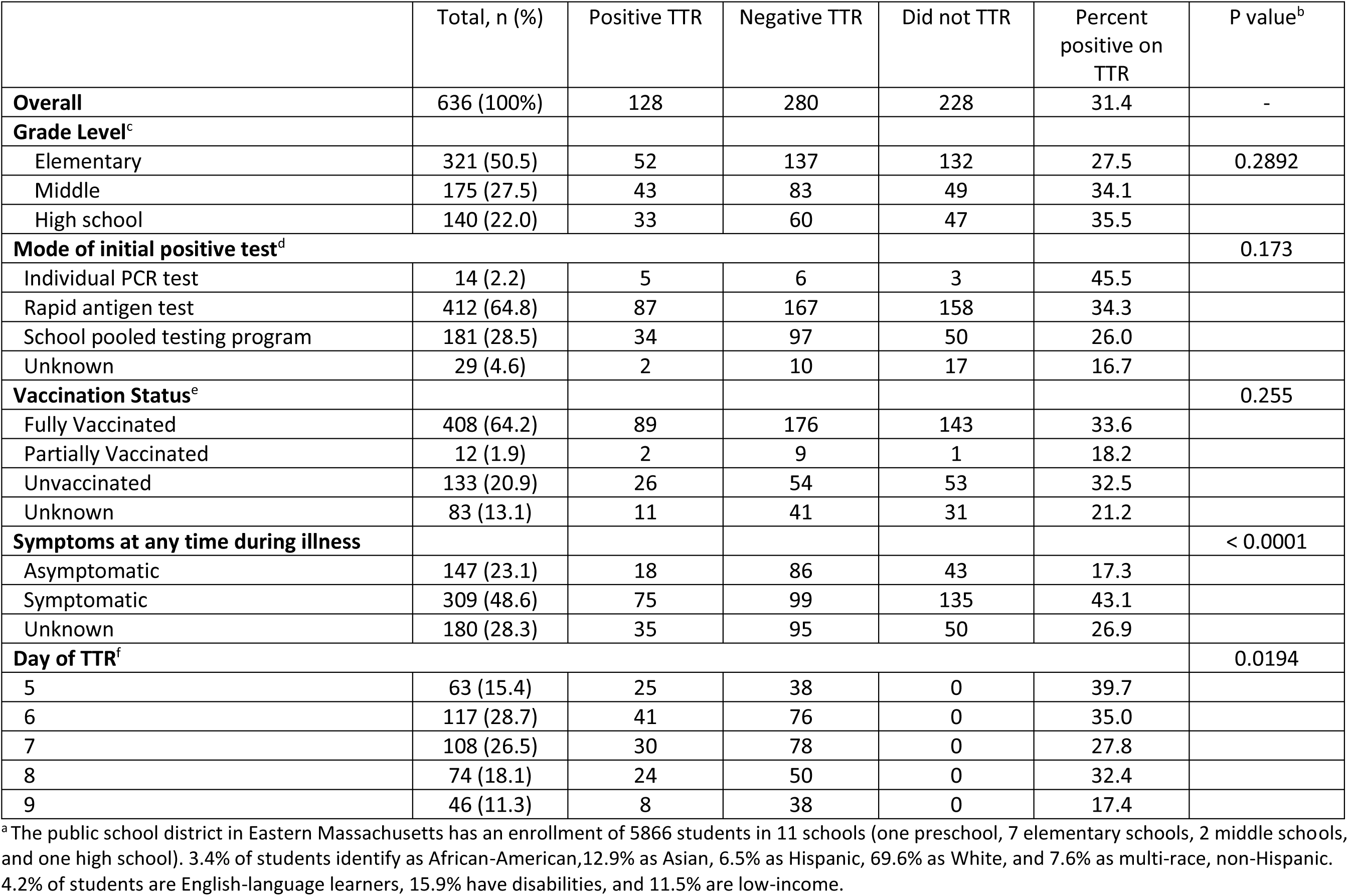
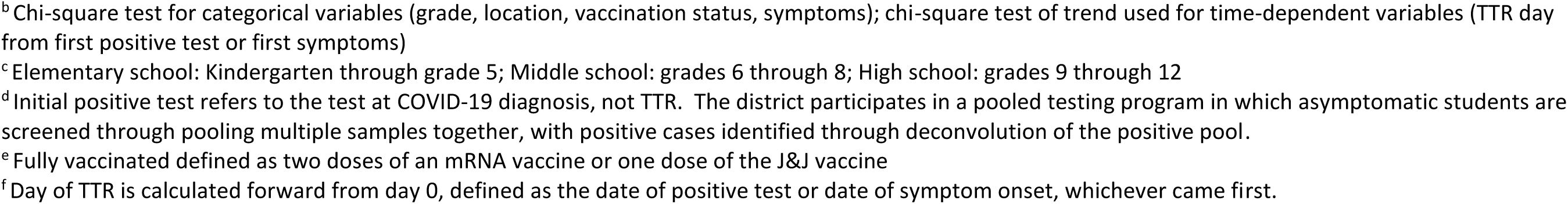
Demographic and clinical characteristics of students with COVID-19 infection eligible for school-based TTR in an Eastern Massachusetts school district^a^

|  | Total, n (%) | Positive TTR | Negative TTR | Did not TTR | Percent positive on TTR | P value<sup>b</sup> |
| --- | --- | --- | --- | --- | --- | --- |
| **Overall** | 636 (100%) | 128 | 280 | 228 | 31.4 | - |
| **Grade Level<sup>c</sup>** |  |  |  |  |  |  |
| Elementary | 321 (50.5) | 52 | 137 | 132 | 27.5 | 0.2892 |
| Middle | 175 (27.5) | 43 | 83 | 49 | 34.1 |  |
| High school | 140 (22.0) | 33 | 60 | 47 | 35.5 |  |
| **Mode of initial positive test<sup>d</sup>** |  |  |  |  |  | 0.173 |
| Individual PCR test | 14 (2.2) | 5 | 6 | 3 | 45.5 |  |
| Rapid antigen test | 412 (64.8) | 87 | 167 | 158 | 34.3 |  |
| School pooled testing program | 181 (28.5) | 34 | 97 | 50 | 26.0 |  |
| Unknown | 29 (4.6) | 2 | 10 | 17 | 16.7 |  |
| **Vaccination Status<sup>e</sup>** |  |  |  |  |  | 0.255 |
| Fully Vaccinated | 408 (64.2) | 89 | 176 | 143 | 33.6 |  |
| Partially Vaccinated | 12 (1.9) | 2 | 9 | 1 | 18.2 |  |
| Unvaccinated | 133 (20.9) | 26 | 54 | 53 | 32.5 |  |
| Unknown | 83 (13.1) | 11 | 41 | 31 | 21.2 |  |
| **Symptoms at any time during illness** |  |  |  |  |  | < 0.0001 |
| Asymptomatic | 147 (23.1) | 18 | 86 | 43 | 17.3 |  |
| Symptomatic | 309 (48.6) | 75 | 99 | 135 | 43.1 |  |
| Unknown | 180 (28.3) | 35 | 95 | 50 | 26.9 |  |
| **Day of TTR<sup>f</sup>** |  |  |  |  |  | 0.0194 |
| 5 | 63 (15.4) | 25 | 38 | 0 | 39.7 |  |
| 6 | 117 (28.7) | 41 | 76 | 0 | 35.0 |  |
| 7 | 108 (26.5) | 30 | 78 | 0 | 27.8 |  |
| 8 | 74 (18.1) | 24 | 50 | 0 | 32.4 |  |
| 9 | 46 (11.3) | 8 | 38 | 0 | 17.4 |  |

<sup>a</sup> The public school district in Eastern Massachusetts has an enrollment of 5866 students in 11 schools (one preschool, 7 elementary schools, 2 middle schools, and one high school). 3.4% of students identify as African-American, 12.9% as Asian, 6.5% as Hispanic, 69.6% as White, and 7.6% as multi-race, non-Hispanic. 4.2% of students are English-language learners, 15.9% have disabilities, and 11.5% are low-income.
<sup>b</sup> Chi-square test for categorical variables (grade, location, vaccination status, symptoms); chi-square test of trend used for time-dependent variables (TTR day from first positive test or first symptoms)
<sup>c</sup> Elementary school: Kindergarten through grade 5; Middle school: grades 6 through 8; High school: grades 9 through 12
<sup>d</sup> Initial positive test refers to the test at COVID-19 diagnosis, not TTR. The district participates in a pooled testing program in which asymptomatic students are screened through pooling multiple samples together, with positive cases identified through deconvolution of the positive pool.
<sup>e</sup> Fully vaccinated defined as two doses of an mRNA vaccine or one dose of the J&J vaccine
<sup>f</sup> Day of TTR is calculated forward from day 0, defined as the date of positive test or date of symptom onset, whichever came first.

**Figure 1.**
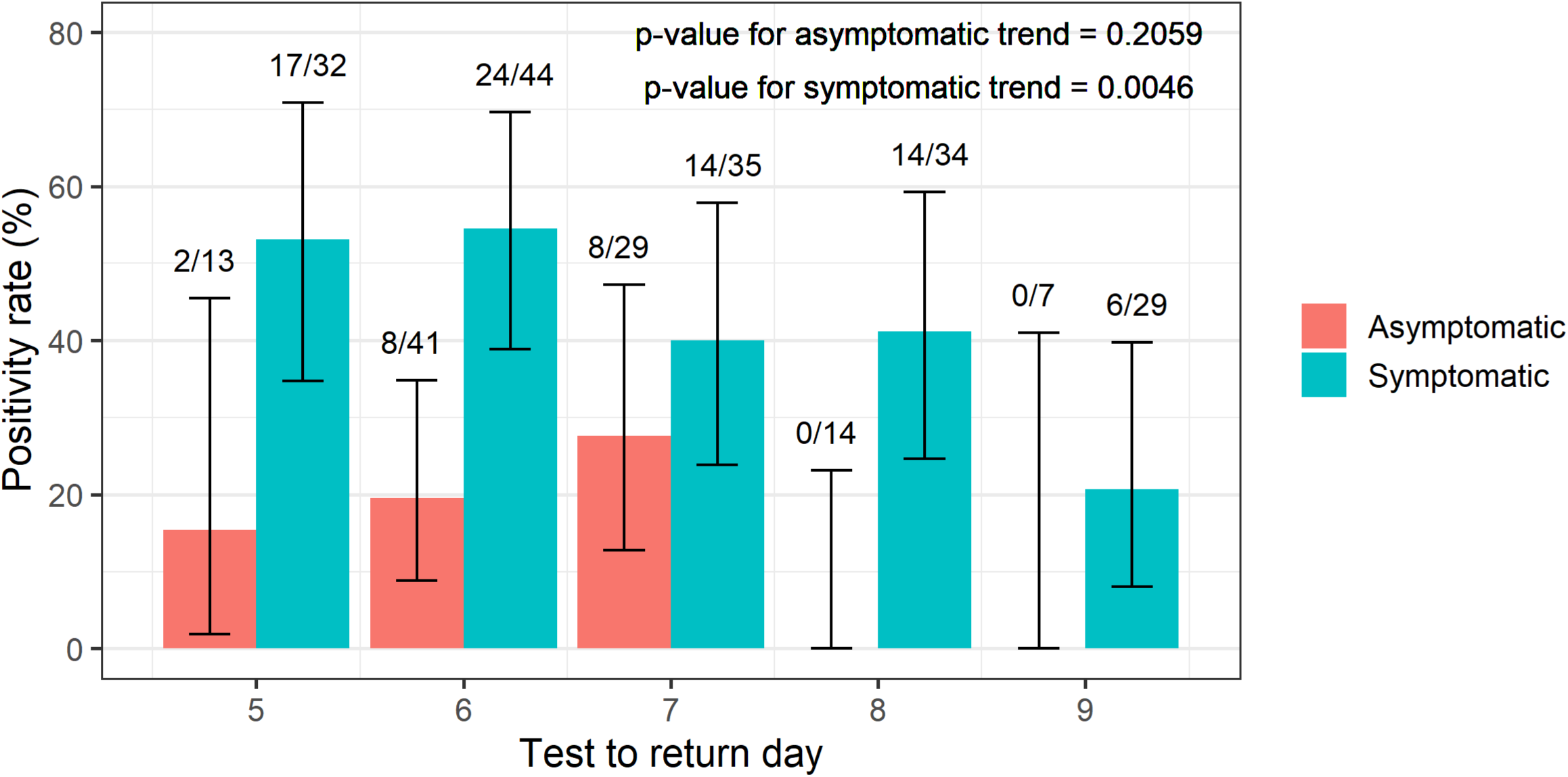
Test to return positivity rate according to day of infection on which TTR was conducted, stratified by presence or absence of symptoms at any time during infection, among those with available symptom status. Chi-square tests of trend were performed for each group. Error bars show 95% exact binomial confidence intervals.

## Discussion

In a public school district during the Omicron surge, 31% of students with SARS-CoV-2 infection had a positive RAT 5-9 days after symptom onset or positive test, despite symptom improvement. This proportion is similar to model predictions^6^ and observed results in health care workers.^7^ Ever-symptomatic students were more likely to have a positive TTR than students who remained asymptomatic (43 vs. 17%).

This study is subject to several limitations. First, students had only one opportunity to participate in TTR; some may have pre-tested or delayed TTR to ensure a negative school-administered test. If all persons meeting time and symptom criteria had tested on day 5 or 6, the positivity rate would likely have been higher. Second, the correlation between RAT and culture-positivity is not yet understood for the Omicron variant, although preliminary data suggests that RAT remains sensitive when viral load is high by RT-PCR.^8^

Implementation of TTR programs in schools has both advantages and costs. Compared to schools that allow return on day 6 without TTR, schools with TTR programs may have fewer school-associated transmissions, but more missed school days. However, as an alternative to isolation through day 10, TTR programs will reduce the number of missed school days. While TTR programs may identify those individuals who still carry replication-competent virus, spread in schools has been rare when masks are worn consistently; further, the risk of school-associated transmission during days 6-10 is not known. When return occurs prior to day 11 without TTR, districts should assume that some students could still be infectious. For students on days 6-10, strict adherence to masking (consistent with CDC guidance) and safe distance during unmasked periods, including lunchtime, are essential.

## Data Availability

All data produced in the present study are available upon reasonable request to the authors and subject to the rules of the collaborating district.

## References

1. Centers for Disease Control and Prevention. CDC Updates and Shortens Recommended Isolation and Quarantine Period for General Population. Published December 27, 2021. Accessed February 9, 2022. https://www.cdc.gov/media/releases/2021/s1227-isolation-quarantine-guidance.html

2. Jefferson T, Spencer EA, Brassey J, Heneghan C. Viral cultures for Coronavirus Disease 2019 infectivity assessment: a systematic review. Clinical Infectious Diseases. 2021;73(11):e3884–e3899.

3. Pekosz A, Parvu V, Li M, et al. Antigen-Based Testing but Not Real-Time Polymerase Chain Reaction Correlates with Severe Acute Respiratory Syndrome Coronavirus 2 Viral Culture. Clinical Infectious Diseases. 2021;73(9):e2861–e2866. doi:10.1093/cid/ciaa1706

4. Chiu C, Killingley B, Mann A, et al. Safety, tolerability and viral kinetics during SARS-CoV-2 human challenge. doi:10.21203/rs.3.rs-1121993/v1

5. R Core Team. R: A language and environment for statistical computing. R Foundation for Statistical Computing, Vienna, Austria. Published online 2021. Accessed February 6, 2022. https://www.R-project.org/

6. UK Health Security Agency. COVID-19 self-isolation changes: scientific summary. Published online 2022. Accessed February 9, 2022. https://ukhsalibrary.koha-ptfs.co.uk/wp-content/uploads/sites/40/2022/01/20220110_Self-isolation_Scientific-Summary_Final-clean.pdf

7. Landon E, Bartlett AH, Marrs R, Guenette C, Weber SG, Mina MJ. High Rates of Rapid Antigen Test Positivity After 5 days of Isolation for COVID-19. doi:10.1101/2022.02.01.22269931

8. Schrom J, Marquez C, Pilarowski G, et al. Title: Direct Comparison of SARS Co-V-2 Nasal RT-PCR and Rapid Antigen Test (BinaxNOW TM) at a Community Testing Site During an Omicron Surge. doi:10.1101/2022.01.08.22268954

